# The Dichoptic Contrast Ordering Test: A method for measuring the depth of binocular imbalance

**DOI:** 10.1101/2024.08.05.24311503

**Authors:** Alex S Baldwin, Marie-Céline Lorenzini, Annabel Wing-Yan Fan, Robert F Hess, Alexandre Reynaud

## Abstract

In binocular vision, the relative strength of the input from the two eyes can have significant functional impact. These inputs are typically balanced, however in some conditions (e.g. amblyopia) one eye will dominate over the other. To quantify imbalances in binocular vision, we have developed the Dichoptic Contrast Ordering Test (DiCOT). Implemented on a tablet device, the program uses rankings of perceived contrast (of dichoptically-presented stimuli) to find a scaling factor that balances the two eyes. We measured how physical interventions (applied to one eye) affect the DiCOT measurement. These were: i) neutral density filters, ii) Bangerter filters, and iii) optical blur introduced by a +3 D lens. The DiCOT results were compared to those from the Dichoptic Letter Test (DLT). Both the DiCOT and the DLT showed excellent test-retest reliability, however the magnitude of the imbalances introduced by the interventions was greater in the DLT. To find consistency between the methods, rescaling the DiCOT results from individual conditions gave good results. However, the adjustments needed for the +3 D lens condition were quite different from those for the ND and Bangerter filter. Our results indicate that the DiCOT and DLT measure partially separate aspects of binocular imbalance. This supports the simultaneous use of both measures in future studies.

## 1 Introduction

When operating under normal healthy conditions, our brains combine the visual input from the two eyes to give a unified binocular percept. This combination process is only partially understood. It is known to involve both excitatory and inhibitory influences between the inputs from the two eyes (Ding and Sperling, 2006; Meese et al., 2006). With basic psychophysics, we can explore these interactions by looking at performance on tasks involving binocular summation and interocular suppression. While these interactions are typically “balanced” (e.g. suppression from the left eye to the right is similar to that from the right eye to the left), they can be severely imbalanced in some visual conditions such as amblyopia (Hess et al., 2014).

An imbalance in binocular vision may be observed due to a weaker input from one eye, reducing its contribution to the binocular percept. This may also have a compounding effect, where that weaker signal also leads to that eye exerting a weaker suppression on the other eye. Alternatively, the two eyes may provide an equal strength of input, with the weight of suppression applied from each eye to the other being imbalanced (Wang et al., 2021). The resulting imbalances can be observed through masking studies, either by measuring interactions between stimuli at corresponding locations in the two eyes (dichoptic overlay masking) or with stimuli at nearby non-overlapping locations (dichoptic surround masking). The two methods have been shown (Huang et al., 2012) to reveal separable aspects of binocular combination and suppression. Even the suppression effects have been shown to be complex, as demonstrated both by the tuning functions for dichoptic masking (Baker and Meese, 2007) and differential effects of short-term monocular deprivation that depend on mask orientation (Baldwin and Hess, 2018). It is important to note that in some measurements it may be difficult to distinguish between a situation where an eye is providing a weaker input and one where an eye is providing an equally strong input but is penalised by greater interocular suppression. For the brief measurements we discuss here the two will likely have equivalent effects.

There can also be a dynamic component to the imbalance between the eyes where the dominance of each eye’s percept varies over time. This phenomenon is studied with the binocular rivalry paradigm, where incompatible spatially-overlapping stimuli seen by the two eyes compete. This results in the percept switching between the two eyes’ inputs (Blake and Camisa, 1979). The binocular rivalry phenomenon is thought to involve a combination of low level (eye-based suppression) and higher-level (perhaps attentional) factors. There is also a further dynamic component of plasticity on rivalry, where perceptual experience can affect the degree of exclusivity of the monocular percepts (Klink et al., 2010). For an expert review of binocular rivalry, the reader is directed to (Alais, 2012). In the current study, we are interested in measuring imbalance in the effective strength of stable monocular percepts, whatever the cause.

While our understanding of the mechanisms underlying binocular vision has advanced in recent years, it is far from complete (Hess et al., 2014). A plethora of laboratory approaches have been devised to quantify imbalances in binocular vision (such as those induced by short-term deprivation of one eye, see various methods compared in Min et al., 2021); a non-exhaustive list is provided in Table A1 of Appendix A. Each depends on a contrast-based metric which is consistent with the current view that the site of interocular suppression is located early in the cortical pathway where contrast gain control occurs (Baker et al., 2008; Meese et al., 2006). These studies have provided information on the factors which influence suppression such as the spatial and temporal properties, and the visual locus. A number of these different laboratory-validated approaches have been simplified to enable their use in the clinic, these include; motion coherence (Black et al., 2011), Dichoptic Letters Test (Kwon et al., 2015), the Worth 4 Dot test app (Webber et al., 2020), phase combination (Ding and Sperling, 2006) and orientation combination (Wang et al., 2019). Of these, it is only the Worth 4 Dot test app that does not require elaborate computer equipment, as it is implemented on a self-contained handheld digital device. It has been shown to be effective in measuring the size of the suppression scotoma, though it may fall short in the measurement of the depth of suppression, possibly due to the added complication of having chromatic as well as achromatic contrast defining the stimuli (Webber et al., 2020).

We have developed the Dichoptic Contrast Ordering Test (DiCOT) as a tool for making quick measurements of the relative contribution of the two eyes to binocular vision. A prototype application has been implemented on a handheld digital device (iPad tablet) for convenience and portability. The subject views the screen through red/green anaglyph glasses, allowing for a dichoptic presentation of stimuli. The principle of the test is that an imbalance between the two eyes (i.e. due to imbalanced suppression) will cause the apparent contrast seen by one eye to be greater than that seen by the other (Bagolini, 1957; Reynaud and Hess, 2016). The goal of the measurement is to find a contrast scaling factor that equalises the contrast seen by the two eyes.

The contrast scaling factor is found by showing a set of images (Figure 1A) to the two eyes of various contrasts (three to the left eye and three to the right eye, in the current version of the method) and asking the participant to rank them in order from most to least “visible” (having previously trained the participant to understand “visibility” to refer to contrast). A single scaling factor adjusts the balance of contrasts seen by the two eyes, searching for a compensation that counter-acts any binocular imbalance. To find the scaling factor a series of “trials” are tested. On each trial, a new set of contrasts are shown to each eye, with the stimulus set selected by an entropy-minimising procedure that allows the tool to efficiently determine the most likely scaling factor in a small number of trials. This is achieved through a look-up table that maps the relative likelihood of each ranking order to a quantised list of possible scaling factors (Figure 1B). Bootstrapping is used to deliver this most likely value along with a 95% confidence interval. An associated measurement error is a fundamental requirement of any clinical test that is involved with therapeutic assessment/endpoint evaluation as it allows statistical evaluations on a single case basis.

**Figure 1:**
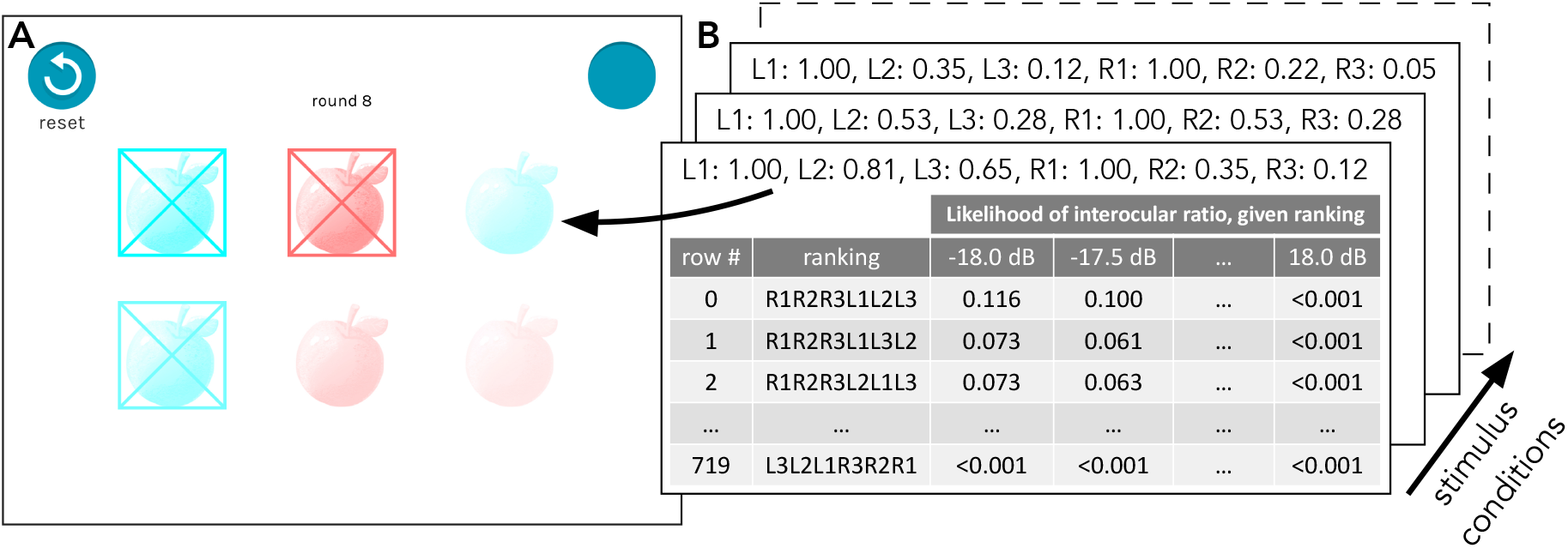
Stimulus display for a single DiCOT trial. Panel **A** shows a screenshot of a trial with an arrangement of left eye stimuli (in red) and right eye stimuli (in cyan) with each set having a high, medium, and low contrast stimulus determined by the algorithm to provide information on the binocular imbalance of the participant. Depending on the degree of imbalance, in some cases the lower contrast stimuli may not be visible to the participant. Panel **B** shows an illustration of the look-up tables (generated by Monte Carlo simulation, one per stimulus condition). Each column is a probability distribution, giving the probability of a particular ranking response being made for that stimulus condition by an individual with that interocular ratio. Each row is a likelihood function, where the ranking made in response to a stimulus condition indicates the likelihood of the participant’s visual system being balanced by some interocular ratio parameter (from −18 to +18 dB).

In the prototype application, there are three small images (cartoon pictures of fruit) shown to each eye. A simpler version of the measurement would present only one image to each eye and simply ask which is of higher apparent contrast, however we have a few reasons for preferring a ranking of multiple images. The first is that on each trial, one of the images (the “high contrast” image) shown to each eye is always at the maximum (100%) contrast. The other two images (“medium” and “low” contrast) vary in their contrast as determined by the algorithm. The goal of the fixed high contrast image is to prevent any eye-based contrast adaptation or renormalisation that might possibly occur over the course of the measurement. The second reason is that the participant implicitly makes multiple comparisons of apparent contrast (between all of the available images on the display) at the same time. This provides more information about the correct scaling from a single presentation. The stimuli are shown continuously until the participant has completed the response for that trial. The outcome of this approach is a simple, game-like measurement involving touching elements on the screen in order of their perceived visibility. This task can be accomplished by children at least as young as 8 years old (Reynaud et al., in review).

A variety of approaches have been evaluated to measure the strength of interocular suppression with a contrast-based metric (see Table A1). However, we would not consider any of these yet as a widely-accepted clinical “gold standard” for the quantitative measurement of the strength of suppression. In this study we therefore choose to compare our test to the recent Dichoptic Letters Test (DLT) (Kwon et al., 2015) because, like ours, it is designed as a clinical tool to quantify the binocular imbalance that may be due to suppression, and it has recently come into common use in the literatur (Birch et al., 2016; Martín et al., 2020; Sauvan et al., 2019). In this study, we introduce the new DiCOT method to measure imbalances in binocular vision. We analyse the test-retest reliability of the DiCOT measurements and compare that result to the DLT. We also compare the results obtained by each measurement to the other. To simulate a range of binocular imbalances, measurements are taken from participants with normal vision, making use of three methods to introduce interocular imbalances: neutral density (ND) filters, Bangerter filters, and optical blur (Li et al., 2012).

## 2 Methods

### 2.1 Participants

The experiments were conducted in accordance with the procedures and protocol approved by the institutional research ethics board of the McGill University Health Center and complying with the Declaration of Helsinki. Written informed consent was obtained. A total of 12 adult participants (age range 19-71 years) took part in this study, with normal or corrected-to-normal vision. This was a number sufficient to estimate test-retest ICC scores for each filter or lens condition with an expected precision of *±*0.1 (Bonett, 2002). Four of the participants were study authors (AR, AWF, MCL, RFH). The remaining four participants were recruited from the local McGill University Health Centre community through word of mouth and posters. They were naïve to the expected impact of the experimental manipulations on their behaviour in the DiCOT task.

All participants were tested wearing their best correction. Screening was performed by the experimenters responsible for the study. Visual acuity was measured using a Snellen chart and stereoacuity was measured with the Randot Preschool Stereotest (Stereo Optical Co. Inc., Chicago, IL, USA). Normal vision was defined as better than or equal to 0.0 logMAR acuity in each eye with stereoacuity < 400 arc sec. Had any participants reported any known visual, cognitive or neurological impairments they would have been excluded, however this was not the case for any of our recruits.

### 2.2 Experiment Design

The relative strength of the two eyes, taken as an index of the imbalance of the two eyes’ contribution to binocular vision, was measured using the existing Dichoptic Letters Test (DLT) (Kwon et al., 2015) and our newly-developed Dichoptic Contrast Ordering Test (DiCOT). Procedures and apparatus for these two tests are detailed below. To produce a wide range of imbalances, measurements were conducted under different lens or filter viewing conditions designed to affect interocular balance. The strength of these manipulations was determined based both on the previous experience of the experimenters and on our initial informal explorations (to try to avoid choosing a penalisation for one eye that could be strong enough to render either task impossible). Vision was deprived by placing either a Bangerter filter (0.8 Power), a 1ND filter (10% transmittance) or a +3 D Lens in front of one eye. Deprivation of both the left eye (OS) and the right eye (OD) was performed for each condition. We also measured the baseline with no filter or lens. This made for a total of 7 conditions to be tested per subject. To measure test-retest reliability, all measures were performed twice in a randomised order.

### 2.3 Dichoptic Contrast Ordering Test (DiCOT)

#### 2.3.1 Stimuli and Apparatus

The DiCOT stimuli were displayed dichoptically using colour anaglyph presentation. Cyan (with contrast only in the Green and Blue colour channels) and red (with contrast only in the Red colour channel) cartoon drawings representing fruit (i.e., apple, cherry or grape) with normalised RMS contrast (normalised for luminance contrast across the different fruits and different anaglyph eye presentations) were displayed on an iPad (iPad mini 5th generation A2133; resolution: 2048 ×1536 pixels, 128 pixels per cm) display with a uniform white background (561 cd/m^2^, measured with an LS-150 Luminance Meter, Konica Minolta Sensing Americas Inc, NJ, USA) with varying contrasts. The software was written in the Unity Engine (Unity Technologies, Copenhagen, Denmark). The screen was viewed through red/green anaglyph glasses (Gulden Ophthalmics, Pennsylvania, USA) at a distance of 40 cm. The fruit pictures each filled a 2.5 cm (3.6 degrees of visual angle, or “deg”) square box, and were positioned in a square grid arranged 4 cm (5.7 deg) apart centre-to-centre.

Through the red filter the screen luminance was 26 cd/m^2^, whereas it was 52 cd/m^2^ through the green filter. Across this relatively modest range of luminances we would expect contrast constancy to hold (Ashraf et al., 2022), were it not to this would result only in a constant bias of the “balance” point of our measurement. Through the red filter, the Cyan stimuli appeared darker than the background. Through the green filter, the Red stimuli appeared darker than the background. A custom-written shader was used to apply luminance gamma correction for linear contrast control, and to balance the contrast seen by the two eyes through the red/green anaglyph presentation. This means that the contrasts we define for our stimuli are the effective luminance contrast after the image has passed through the coloured anaglyph filter. Spectral measurements from the iPad mini display made with a VPixx-Edition X-Rite i1Pro (VPixx Technologies, Saint-Bruno, Canada) showed that the red and green colour channels were not entirely independent. Attempts to show green stimuli also introduced a small red component. The result of this for our anaglyph presentation was a cross-talk image at 20% of the presented contrast when viewing through the red filter as compared to only 3% contrast when viewing through the green filter. The expected outcome of this cross-talk would be a small bias favouring the green-filter eye.

#### 2.3.2 Procedure

Testing began with a few example pages where participants had to select some binocularly-presented cartoon images of fruit in order from high to low contrast (though the on-screen instructions used the term “visibility”, the examples provided served the purpose of linking the idea of visibility to contrast). Selecting the fruit in the correct order (by tapping the screen with a finger) was a requirement to pass through this training, which ensured participants understood how to perform the task required. The participant then proceeded to the measurement trials.

On each measurement trial, participants saw six cartoon images of fruit (Figure 1A) at various contrasts. These were presented dichoptically so three were seen by each eye. Their task was to select the fruits in order from the highest to lowest apparent contrast by tapping on them with a finger (“Please tap fruits from most to least visible”). When a fruit was tapped there was both visual feedback (the fruit became crossed-out) and auditory feedback. If the participant decided they made a mistake and tapped the wrong fruit they could choose to reset the trial by tapping a “reset” button. The trial completed when all six fruits had been tapped, initiating a 0.5 second delay before the next trial was presented. There is a possibility that on some trials the lower contrast images may not be visible (for example if a measurement was being made under conditions that heavily affected the strength of the input from one eye). Provided the participant had at least one eye with good contrast sensitivity we would always expect a minimum of three fruits (seen by that better eye) to be visible. If any fruits are still not visible the participant is instructed to guess. To prevent the measurement stalling if the participant will not guess, a skip button becomes available 15 seconds after the third fruit is selected. This has the effect of selecting the remaining fruit in random order on behalf of the participant (similar to the “guess button” introduced by García-Pérez, 2010), thus ending the trial.

For the first trial, the stimulus set showed the same contrast range to the two eyes (100%, 53%, 28%). After this, the contrast on each trial was set by an entropy-minimising procedure that attempted to determine the best value for the interocular contrast ratio in the fewest trials. Interpretation of the rankings provided by the participant depends on a set of look-up tables that give the probability of different ranking responses for the different stimulus conditions under various assumed interocular contrast gain ratios (Figure 1B). These were generated through Monte-Carlo simulation (100,000 simulations per stimulus condition and per parameter value which stepped from −18 dB to +18 dB in 0.5 dB intervals). In the simulations, the input gain of the two eyes varies according to the interocular gain ratio, with the judgement of contrast being linear but affected by a 10% internal additive Gaussian contrast “noise”. Varying this noise parameter has the effect of making the likelihood functions overall broader (for large values) or narrower (for small values). The value of 10% was not selected on the assumption that it is a good estimate of the amount of noise affecting this process in human vision. Instead, it is a value which gives reasonably broad likelihood functions that do not challenge the precision limit for representing and manipulating probability values (which may become very small) in the software. In the stimulation, the participant simply selects the targets in order of the amplitude of the noisy response to each target. In the tables, each column gives the relative frequency of the 720 (6! for the ranking of six targets) possible ranking outcomes generated on the basis of a specific stimulus condition and interocular gain ratio. Therefore, the values in each column must add up to 1.

When a participant completes their ranking in response to the stimulus condition, the corresponding row in the relevant look-up table is selected (Figure 2A). This indicates the relative likelihood of each of the possible (quantised) parameter values on the basis of the ranking given in that trial. We say relative likelihood here, as we are ignoring the possibility of the parameter taking a value either between those we sampled (our quantisation was chosen to give a precision which would be realistic based on the length of the measurement) or outside that range (for such large imbalances the algorithm will return its maximum of either −18 dB or +18 dB). The log of this value is added to the sum of all of the previous log likelihoods (Figure 2B) to obtain an overall log-likelihood function, the peak of which indicates the most likely parameter value given the data (under the assumption that the prior for this parameter is flat). A 1% probability of lapsing (responding randomly) on each trial was also incorporated in this calculation. After each trial, the updated likelihoods of the different parameter values were used to select the stimulus condition for the next trial. The selected stimulus condition was the one which minimised the expected posterior entropy from that trial’s response (see Appendix B). In other words, the algorithm considers all of the possible conditions that could be tested on the next trial and (based on the previous responses) chooses the one that will be most informative regarding the imbalance.

**Figure 2:**
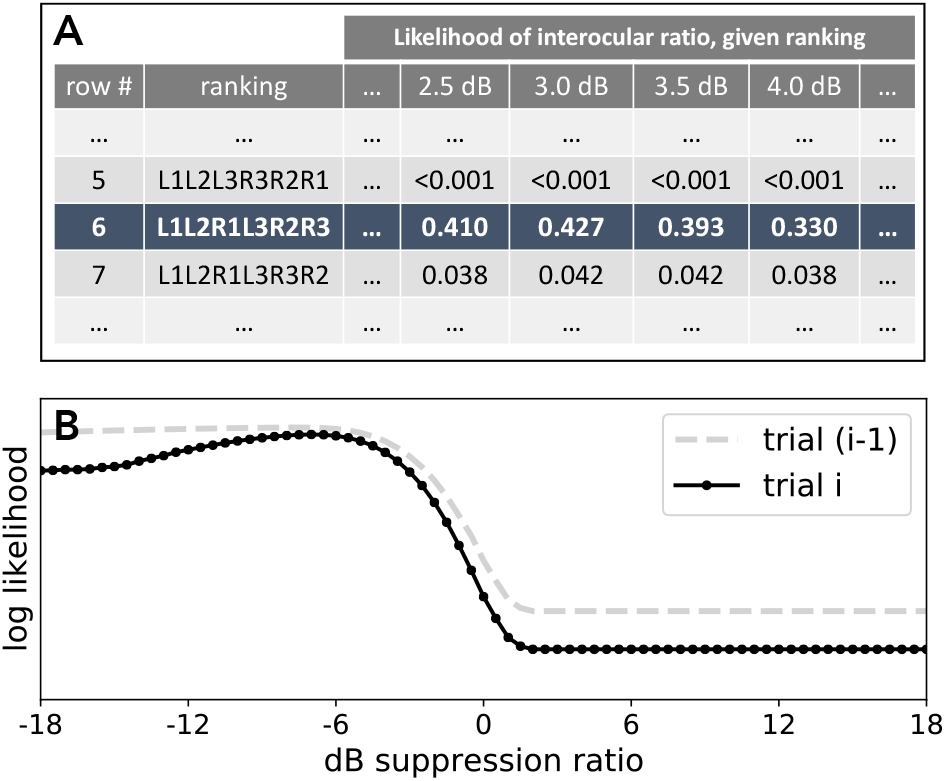
Post-response analysis performed in the DiCOT. The ranking given by the participant selects a row in the look-up table for that trial (**A**). The likelihoods from this row are converted to log values before being added to the cumulative history of the log likelihoods from all previous trials (**B**). The curves in this illustration show relative log likelihoods of the different parameter values based on the cumulative history up to either trial (i - 1), in grey, or up to trial i, in dark blue. This example demonstrates the possible impact of adding trial i, which is to sharpen the estimate of the best parameter value, as well as to vertically translate the function downward.

The algorithm terminated once a certain confidence interval limit (3 dB) was reached (after a minimum of 10 trials) or after a maximum of 20 trials if the confidence intervals were not sufficiently narrow by that time. The 95% confidence interval is determined through non-parametric bootstrapping (1000 samples), randomly resampling from the individual trials and selecting the most likely value from each bootstrapped dataset. A subset of the measurements was timed, from these the typical duration was approximately 2–3 minutes.

### 2.4 Dichoptic Letters Test

#### 2.4.1 Stimuli and Apparatus

The Dichoptic Letters Test was adapted from Kwon et al. (Birch et al., 2016; Kwon et al., 2015). This method consists of reading dichoptically displayed band-pass 2 cycle/deg filtered Sloan letters arranged in a layout similar to the ETDRS acuity chart on a gray background (60 cd/m^2^). The stimuli were generated and controlled using MATLAB (version 9.7.0) with Psychophysics Toolbox extensions for Windows 10, running on a PC desktop computer. Stimuli were presented dichoptically on an active 3D display setup using the Nvidia 3D Vision 2 shutter glasses (nVidia Corp., Santa Clara, CA) running at 60 Hz frame rate per eye and an LCD monitor (model: Asus VS278H-E; refresh rate: 120 Hz; resolution: 1920 ×1080; maximum luminance: 250 cd/m^2^). Participants were seated 50 cm away from the screen in a dimly-lit room. Two different sets of 5 letters of different contrast were presented dichoptically overlapping to each eye. At each position, the identity and interocular contrast-ratio of the letter on each chart differed.

#### 2.4.2 Procedure

The balance point between the two eyes, the interocular contrast-ratio required to report the letter in each eye with equal probability was determined via an adaptive procedure described in detail by Kwon et al. (2015). In each trial, participants were instructed to read aloud the more visible letter at each of the 5 positions in the chart from left-to-right, then the experimenter manually input the letters. The relative contrast of overlapping letters in each eye was adjusted via a customised adaptive procedure adapted from the QUEST (Watson and Pelli, 1983) for a total of 60 data points (12 charts × 5 contrast ratios) (Kwon et al., 2015). The task typically took 1-2 minutes.

### 2.5 Statistical Analysis

Different filters and lenses (i.e., ND filter, Bangerter filter, and blurring lens +3 D, see Methods) were tested twice separately on each eye of the participants performing both tests. In each test, the interocular suppression ratio was expressed in decibels

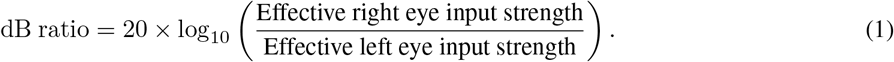

A measurement of 0 dB would indicate perfect binocular balance, with each 6 dB step above or below that equivalent to a factor of 2 ratio in either direction. Penalising the input from the left eye is therefore expected to have the effect of making the dB ratio more positive.

Statistical analysis was performed in R (R Core Team, 2023), with additional packages providing Intraclass Correlation Coefficients (Gamer et al., 2019) and Bland-Altman (Datta, 2017) analysis. We assessed the test-retest reliability of each method using a) the agreement version of the Intraclass Correlation Coefficient (ICC) and b) a Bland-Altman plot (Bland and Altman, 1999) where the difference between the two values (test-retest) is plotted as a function of the mean of the two values for each subject. We then compared the DiCOT against the DLT. When comparing the two tests against each other the consistency version of the ICC was used. We also looked for the best linear relationship between the dB values obtained on the two tests and used this to perform a transformation of the DiCOT values to put them in their best agreement. The relationship between these values and the DLT were also assessed through a scatterplot and a Bland-Altman plot (taking the DLT as the reference value for the x axis).

## 3 Results

### 3.1 Influence of simulated monocular visual loss on interocular suppression ratios

In total, 168 measurements were completed with the DiCOT. For 15 of these measurements the information regarding the confidence interval width was lost. For the remaining 153 measurements, we found that the 3 dB confidence interval width criterion was satisfied in 93% of measurements. The remaining 7% reached 20 trials without the CI width reaching 3 dB or lower. For the DLT there were two participants who in a +3 D Lens condition could not perform the measurement on either the test or the retest measure. For the DiCOT, we looked for a relationship between the absolute degree of imbalance and the associated error (95% confidence interval width). We found no significant relationship (Pearson R = 0.00, P = 0.98).

In Figure 3, we compare the imbalance that was introduced in the results of the two tests for the various filter and lens manipulations we used. Each method introduced an imbalance that could be quantified by both the DiCOT and DLT. These imbalances tended to result in more extreme results on the DLT than on the DiCOT (comparing the standard deviation of all measurements pooled together: 3 dB vs. 10 dB). Figure 3A presents the average measurements for the 12 participants obtained with the DiCOT as a function of the type of filter worn. Deprivation of the right eye would lead to negative values whereas deprivation of the left eye would lead positive values. Figure 3B displays the same measurements but obtained using the Dichoptic Letters Test (DLT). A similar pattern is observed; however, the effect of the lens (+3 D) was more exaggerated in the DLT compared to the DiCOT. We believe this is due to the DLT being tested at a further distance than the DiCOT (50 vs. 40 cm). At this distance, the lens has a more dramatic effect on visual acuity (20/125 reduction compared to 20/50).

**Figure 3:**
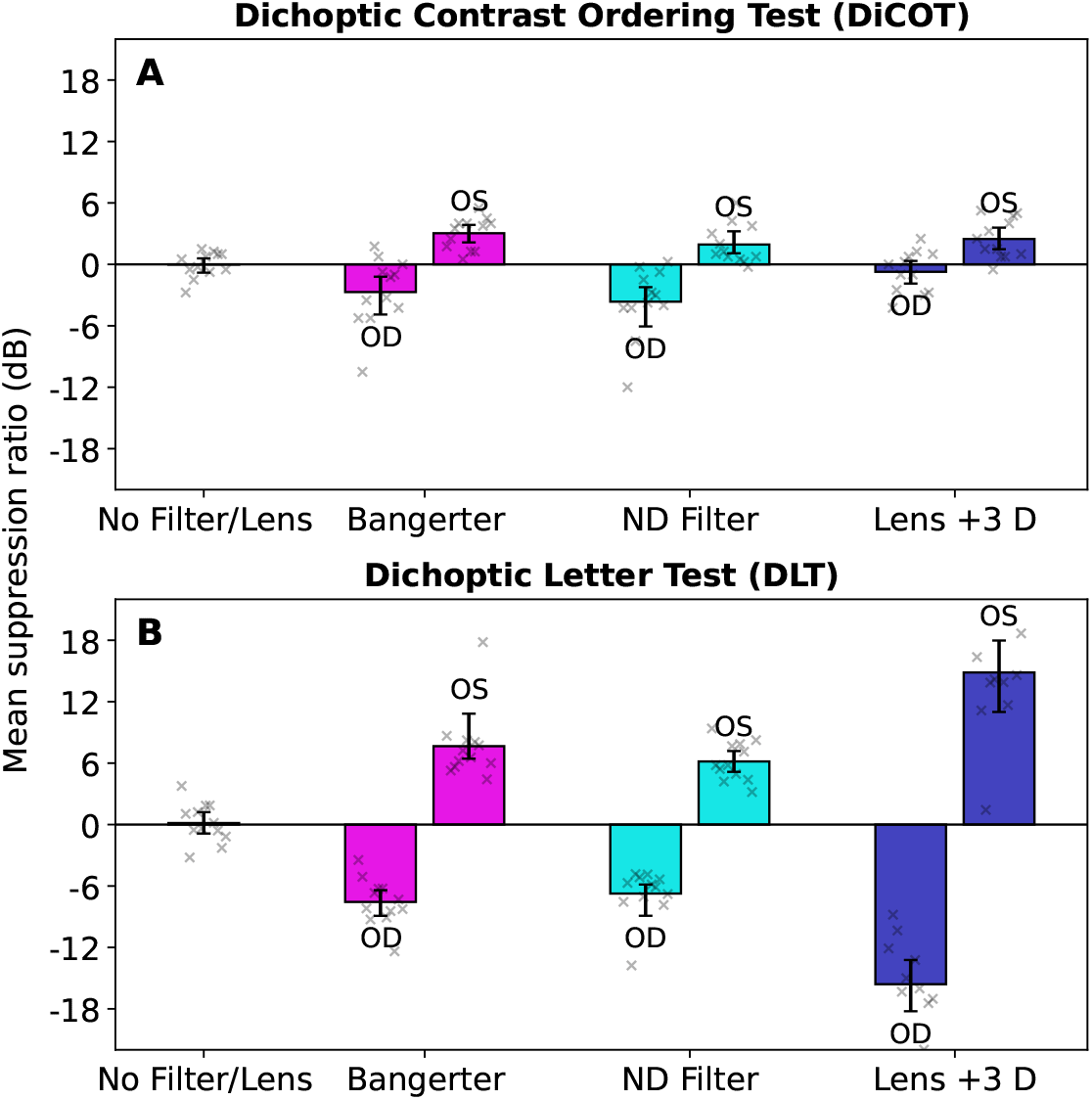
Binocular imbalance without lens/filter and under three lens/filter conditions. The bar graphs represent the mean interocular suppression ratio parameter at baseline and in the 3 conditions where the filter or lens is placed over either the left (OS) or right eye (OD). Panel **A** shows the results from the DiCOT, panel **B** from the DLT. Error bars represent the bootstrapped 95% confidence interval of the mean. Individual participant data are shown for each bar with the cross symbols.

### 3.2 Test-retest reliability of the interocular suppression assessment methods

The test-retest comparisons made with the two measurement tools are shown in Figure 4. The plots in the left column are the test-retest scatter plots. Figure 4A shows this for the DiCOT, with the agreement between the test and retest measurements quantified by the ICC_agreement_ score. This is a specific test of the extent to which the second measurement differs from the first (i.e., it is not a simple correlation measure looking for an unconstrained linear relationship between the two). With this, we obtain a value of 0.91 which can be interpreted as excellent (Cicchetti, 1994). A marginally better result is found for the DLT, shown in Figure 4C (0.97).

**Figure 4:**
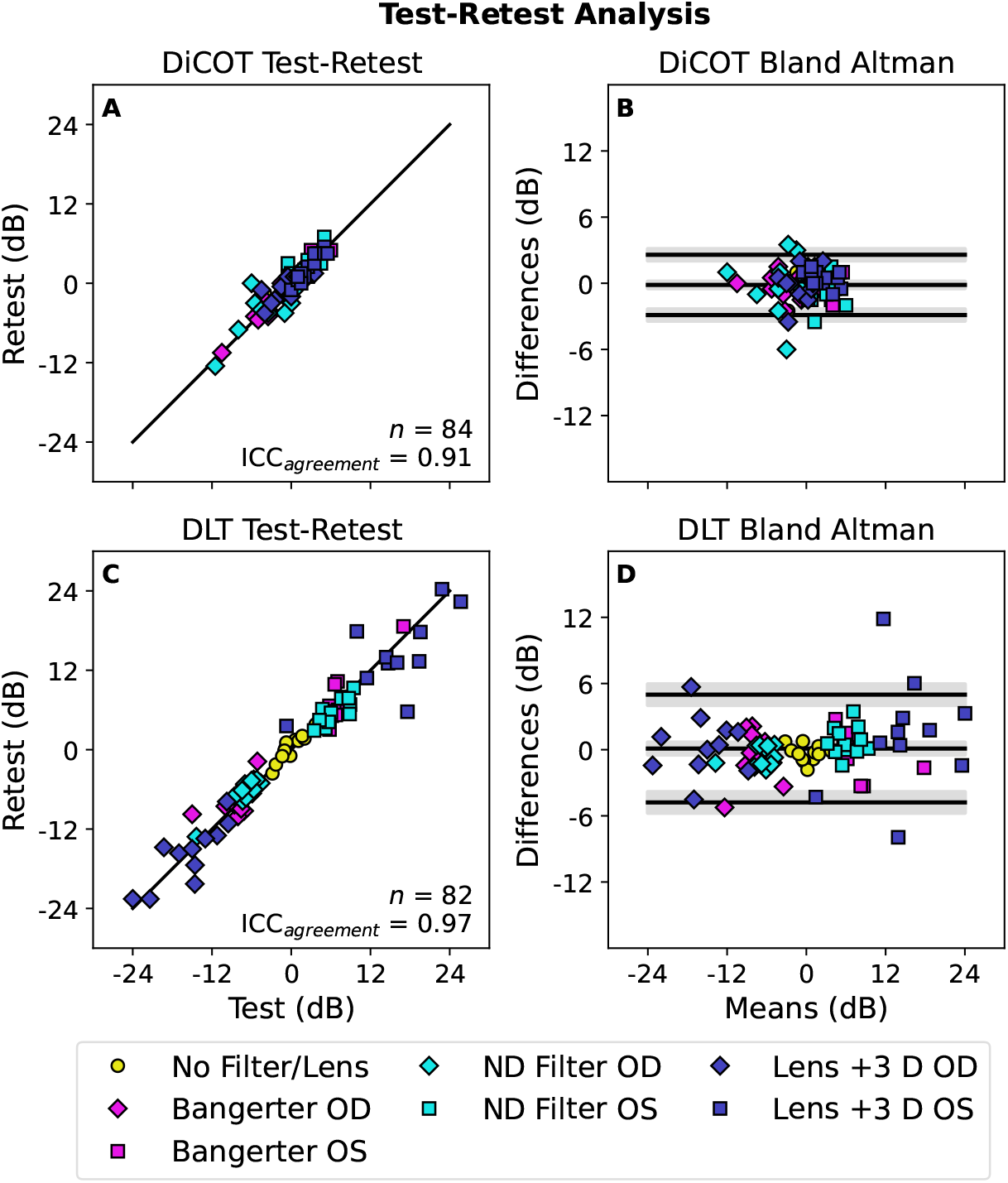
Test-retest reliability. The left column shows correlations between first and second (test and retest) measure- ments obtained using **A** the DiCOT and **C** the DLT. The solid diagonal line indicates equality. Each point indicates a data point from one participant in one lens/filter condition (84 in total for the DiCOT, 82 in the DLT due to cases where a measurement result could not be obtained). The right column shows Bland-Altman plots giving the test-retest difference as a function of the mean of the two measures. The three black lines show the bias and the upper and lower limits of agreement. The shaded region around each line gives the 95% confidence interval. The results for each filter or lens condition are shown individually in Appendix Figure A1, with statistics reported in Table 1.

The right column of Figure 4 shows the Bland-Altman plots for each method. We see that the limits of agreement are narrower (around half the width, see bottom row of Table 1) for the DiCOT than they are for the DLT. This is unsurprising, as the range of values obtained is also narrower for this measurement. Visual inspection reveals no evident systematic relationship between the degree of imbalance and the difference between test and retest measurements.

**Table 1:** Test-retest reliability. Reporting test-retest reliability statistics shown in Figure 4 and Appendix Figure A1. The agreement form of the Intraclass Correlation Coefficient is reported, as well as the bias and the limits of agreement from the Bland-Altman plots.

|  | DiCOT |  | DLT |  |
| --- | --- | --- | --- | --- |
| Condition | ICC<sub>agreement</sub> | Bland-Altman Bias [LOA] | ICC<sub>agreement</sub> | Bland-Altman Bias [LOA] |
| No Filter/Lens | 0.78 | -0.1 [-1.9, 1.7] | 0.92 | -0.3 [-1.8, 1.1] |
| Bangerter | 0.97 | -0.2 [-2.2, 1.7] | 0.97 | -0.4 [-4.3, 3.6] |
| ND Filter | 0.88 | -0.3 [-4.2, 3.6] | 0.98 | +0.1 [-2.4, 2.6] |
| Lens +3 D | 0.89 | +0.1 [-2.3, 2.5] | 0.97 | +0.9 [-7.1, 8.8] |
| **Pooled** | **0.91** | **-0.1 [-2.9, 2.6]** | **0.97** | **+0.1 [-4.8, 5.0]** |

A further analysis was performed on the data from the individual filter and lens conditions. The scatter and Bland-Altman plots are provided in Appendix Figure A1, with the statistics reported in Table 1. In most cases, the test-retest reliability was comparable between the different conditions. The exception was for testing without a filter or lens. For the DiCOT in particular, the ICC_agreement_ score was lower in this condition (0.78). The reason for the poorer performance of the DiCOT in this case is likely due to the smaller range of imbalances found in our study participant population (in the absence of filter or lens manipulations designed to introduce imbalances).

### 3.3 Comparison between interocular suppression assessment methods

Interocular suppression ratios obtained with the DLT are compared against those measured with the DiCOT in Figure 5. As in Figure 4, each participant and lens/filter condition is plotted as a separate point. The top and bottom rows give the data from the test and retest measurements respectively. The consistency ICC between the two measurements was poor (0.32) in both cases. Looking for any linear relationship in the scatter plots (left column), we find best-fitting lines with slopes of 0.18 on the log-log axes for both the test and retest measurements. This accounts for 27-29% of the variance. Because the slope of this relationship was not unity, we find a systematic relationship between the DLT result and the difference between the DLT and DiCOT results in the Bland-Altman plots (right column of Figure 5).

**Figure 5:**
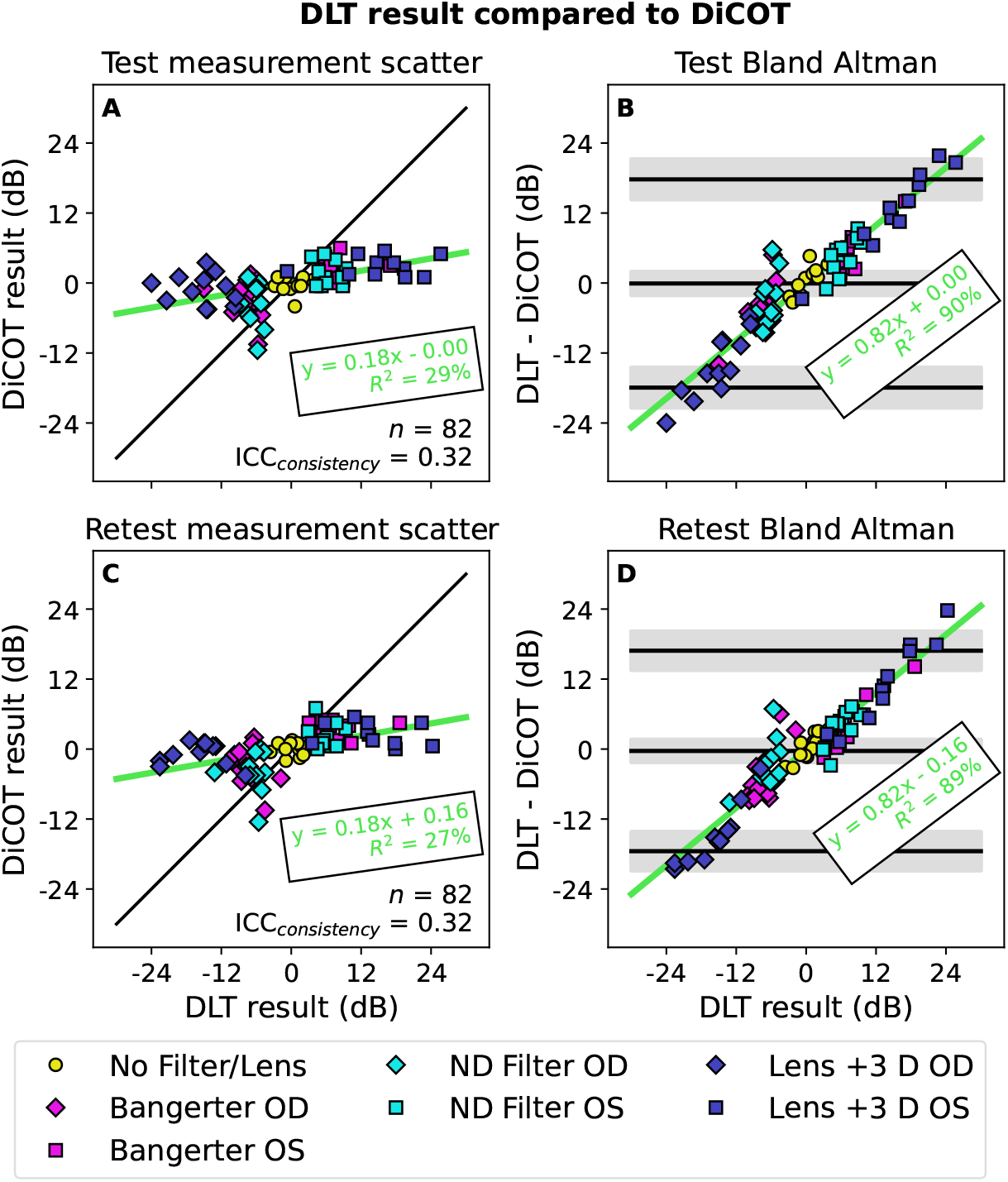
Comparison between DLT and DiCOT. The left column shows scatterplots comparing the measurement made with one test against that made with the other. The top row compares the test (first) measurements, and the bottom compares the second (retest measurements). The diagonal green line is the best-fitting linear relationship (parameters given in green text). The black line shows equality. The right column shows Bland-Altman plots derived from the scatter plots. The three black lines show the bias and the upper and lower limits of agreement. The shaded region around each line gives the 95% confidence interval. The diagonal green line is the best-fitting linear relationship (parameters given in green text). The results for each filter or lens condition are shown individually in Appendix Figure A2, with statistics reported in-part in Table 2 and in-full in Appendix Table A1.

We performed a separate analysis of the relationship between the DiCOT and DLT results obtained with the different filter and lens manipulations. This is plotted in Appendix Figure A2 with the full statistics reported in Appendix Table A1. The ICC_consistency_ was poorest for the measurements without filter or lens, likely due to the small baseline variation in the imbalance in our population relative to the variability in the measurements. The ICC_consistency_ for the Lens +3 D condition was also lower than for the other filter and lens conditions, as would be expected based on the differences in the magnitudes of the effects seen in Figure 3. These magnitudes would affect the slope of the linear relationship between the DLT and the DiCOT, which the ICC_consistency_ metric is sensitive to (as it assumes that the effects of the different measurements on the two tests will have equal magnitude and so the slope will be unity). The slopes from the scatter plots are summarised in Table 2. Without filter or lens there was no significant relationship between the DiCOT and the DLT. The Bangerter and ND Filter conditions had a steeper slope (0.31 to 0.39) whereas the Lens +3 D condition had a shallower slope (0.10).

**Table 2:**
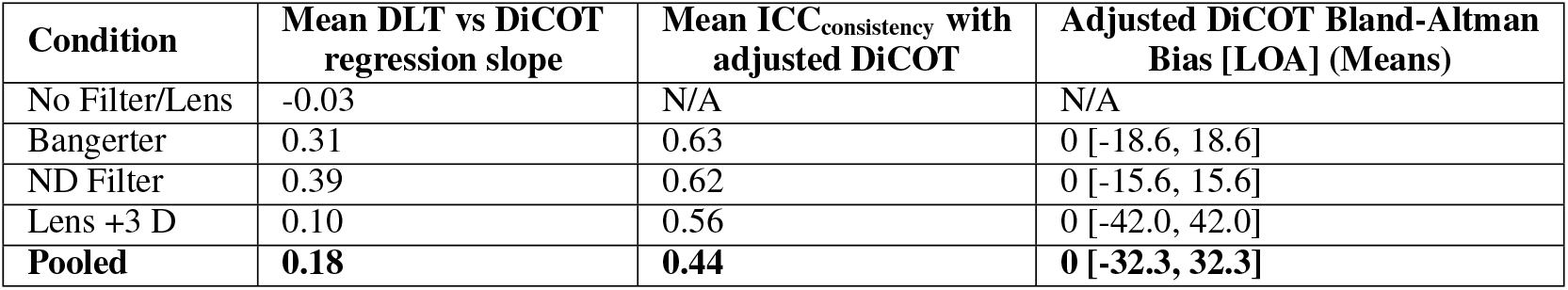
ICC consistency calculated for the individual lens/filter conditions (as shown in Figure 6), in each case combining the OS and OD data. As there was no significant linear relationship for the No Filter/Lens condition, they were not included for the individual analysis with the Adjusted DiCOT.

To improve the consistency between the DLT and DiCOT, we used the best-fitting linear slope and constant parameters to transform the DiCOT results such that they should have their best correlation with the DLT. The relationships with the adjusted DiCOT results are shown in Figure 6. As expected, this slightly improves the ICC to 0.43–0.45 (“fair” by Cicchetti, 1994). The right column shows the Bland-Altman plots calculated on the basis of the adjusted DiCOT values. The limits of agreement here are very broad (*±* 32 dB). There are a variety of differences between the two measurements which may account for the discrepancy between the results obtained with them. We performed an additional, separate analysis for each of the filter and lens conditions. These were transformed based on the linear relationships for each condition reported in Appendix Table A1. As there was no significant relationship for the No Filter/Lens condition, that condition was excluded from this analysis.

**Figure 6:**
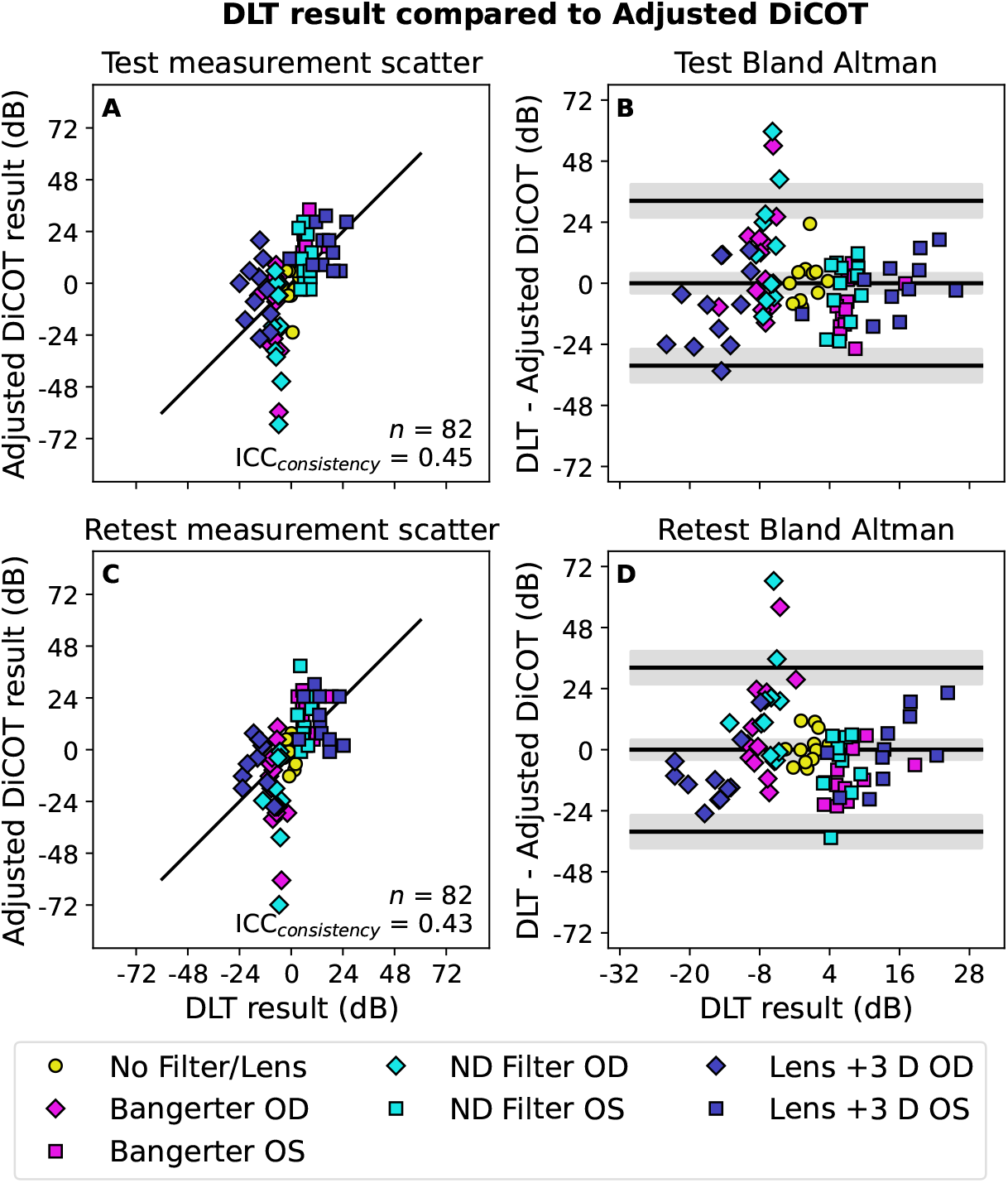
Comparison between DLT and adjusted DiCOT measurements. Similar to Figure 5, however in this case the DiCOT values are adjusted according to the linear relationships reported in Figure 5A (for the test measurements) and Figure 5C (for the retest measurements). The results for each filter or lens condition are shown individually in Appendix Figure A3, with statistics reported in-part in Table 2 and in-full in Appendix Table A2.

The mean ICC_consistency_ for each filter or lens condition is provided in Table 2. In each case, the consistency between the adjusted DiCOT and DLT values was better (0.56 to 0.63) for the individual conditions than for the pooled data (0.44). So overall, we do see consistency between the two tests within an individual lens or filter condition, but the relative scaling of the impact of the different manipulations varies between the two methods. Specifically, the Bangerter filter and ND filter seem to have a similar relationship, but the +3 D Lens has a relatively small effect measured in the DiCOT compared to the DLT (as shown in Figure 3). This may be expected, as the viewing distance for the DLT was greater and so the impact of the blur from the lens would also be greater. We also see much larger Bland-Altman limits of agreement in this condition compared to the Bangerter or ND Filter conditions.

## 4 Discussion

This study introduces a new quick method, the Dichoptic Contrast Ordering Test (DiCOT), for measuring the relative strength of the two eyes’ contribution to binocular vision. With anaglyph presentation, it can be delivered on a variety of digital devices. It involves a simple task of ranking the visibility of dichoptically-presented stimuli, which after only 10–20 trials provides a quantitative estimate of the magnitude and direction of any imbalance along with the associated measurement error. It has excellent test-retest reliability both in the combined analysis of all of our conditions (Figure 4A) and in the analysis of the results from individual filter or lens conditions (Appendix Figure A1).

We chose the Dichoptic Letters Test (DLT) (Kwon et al., 2015) to compare its results with those obtained from the DiCOT. We do not expect that the two methods measure an identical interocular suppression process. The DLT measures overlay suppression, namely the suppression between corresponding regions of the visual field in both eyes which are presented with rivalrous stimuli. Our DiCOT on the other hand measures dichoptic differences in perceived contrast, which may be mediated by suppression arising from surrounding regions of the visual field from the other eye. The DLT result we expect to mostly reflect feedforward and local inter-cortical interactions while the DiCOT would be expected to depend more on feedback interactions from higher cortical areas (Angelucci and Shushruth, 2013). However, what the tests have in common is that they both are suitable to be used in clinical measurements. They are appropriate for children as well as for adult patients, and they both provide the error associated with the measurement.

When penalising with lenses and filters, both tests show changes in the binocular imbalance (Figure 3). The (DLT), which we propose is based on effects of overlay suppression and rivalry dominance (where that rivalry state itself may affect the behaviour of the system, see Klink et al., 2010), gave more extreme values than the DiCOT. Our DiCOT measure is designed to reflect both the feed-forward contrast gain of the two eyes inputs, as well as any effects of dichoptic surround suppression from the elements seen in the other eye. Both tests exhibited excellent test-retest reliability. Although the pattern of results found with the two tests was similar, the results from one accounted for less than half of the variance in the results from the other (Figure 5, Figure A2). We argue that they target (at least partially) distinct aspects of binocular vision and so are differentially affected by our different manipulations. This is reflected by the different regression slopes reported in Table 2. For example, to the extent that the DLT is a rivalry-based task there is an expectation that a blurred input in one eye (from our +3 D Lens) would greatly reduce the dominance of that blurred image (Arnold et al., 2007). On the other hand, we would also expect a (less dramatic) reduction in the perceived contrast in the DiCOT if the image were to be blurred in one eye (May and Georgeson, 2007). This exaggerated effect of blurring on the DLT is reflected visually in our results (Figure 3) and by the relatively shallow regression slope (Table 2). Determining the best instructions and training to separate blur from contrast judgements in the DiCOT is a topic for future research. If we adjust the data from each filter or lens condition individually, we do find they each show a comparable consistency between the DiCOT and the DLT (ICC_consistency_ scores of 0.56 to 0.63 in Table 2). This is still lower than the test-retest reliability, indicating a residual difference between the two measurements. The possibility that the DiCOT and the DLT may target distinct aspects of binocular imbalance makes the combined use of both tests interesting. One such study has been performed comparing the two measures in amblyopia (Reynaud et al., in review).

Although both the DLT and the DiCOT should reflect imbalances in the gain of the input from each eye, we do find a wider range in the DLT results. This may be due to the rivalrous stimulus design. Exaggeration of the imbalance for measurements made with rivalrous stimuli would also be expected in amblyopia (Meier et al., 2023). The other difference of interest to us would be the ways that the two tasks would be affected by interocular suppression. Spatially, suppression can be induced both by stimuli occupying a corresponding retinal locus (overlay suppression) and those in neighbouring regions of the visual field (surround suppression). Overlay suppression is thought to mostly involve local responses from neurons responding within the stimulus area and exhibits orientation tuning. Surround suppression involves more remote suppressive interactions. These involve both intracortical horizontal connections and likely feedback contributions from higher visual areas (Angelucci and Shushruth, 2013). Very remote surround interactions do not exhibit orientational tuning (Shushruth et al., 2013). Interocular suppression in individuals with normal healthy vision that have induced interocular imbalances, or in amblyopic individuals who have intrinsic interocular imbalances as a result of a disruption of normal visual development, involves contributions of both overlay and surround suppression (Huang et al., 2012). This imbalance in amblyopia does not affect behaviour on contrast matching tasks when stimuli are presented monocularly (Hess and Bradley, 1980), suggesting that there is no suprathreshold impact of interocular suppression on perceived contrast under monocular stimulation. Under dichoptic viewing however, contrast matching is impacted (Reynaud and Hess, 2016). The specificity of this phenomenon to simultaneous dichoptic presentation underlies our argument that imbalances measured with the DiCOT can be considered to reflect interocular suppression. However, for many purposes to which this tool may be used, the distinction between imbalances in gain and those in suppression may be irrelevant.

### 4.1 Limitations and Future Work

A number of possible improvements to the DiCOT measurement have been recognised throughout the study process. From the beginning, we have anticipated that the multiple comparisons made by the participant should allow, in principle, for the participants’ responses to be validated for internal consistency. This will be implemented in a future iteration. In the Monte Carlo simulations that generate our look-up tables, we use a linear response to contrast. This means that in the simulated rankings, equal differences in linear contrast are equally discriminable. To better correspond to the human visual system, the look-up tables would be improved if they instead operated on a logarithmic contrast scale. This would make the discriminable differences follow the Weber-Fechner Law where the discriminable increment in some property (such as contrast) is proportional to the baseline amount (Wixted, 2019). The effect of such a change will be to rebalance the relative importance of transpositions made in the ranking of contrasts in the lower or higher contrast range. A linear simulation is “better” at ranking the higher contrast stimuli accurately than a simulation following the Weber-Fechner Law would be. This means that deviations from the expected ranking order made with those higher contrasts are less probable, and so will have a greater impact on the predicted imbalance in the current form of the DiCOT. The effect of changing this rule will be explored in a future study with a new version of the DiCOT.

There is also a need to investigate how technical aspects of the stimulus presentation (e.g. alternatives to the anaglyph mode of dichoptic presentation) and design (spacing between the targets, broadband vs. spatially bandpass targets) affect the measurements. In the current study, the DiCOT presented via red/green anaglyph on an iPad is compared against the DLT presented on a 3D display system using active shutter glasses. A comparison was made between the two measurements on the different technologies as there was not a version of the DLT available for colour anaglyph, nor do we have a version of the DiCOT for an active 3D system. So, effects due to the equipment may have impacted any differences we found. A comprehensive analysis of these factors will be pursued in future studies. The participants free-viewed the stimuli without eye-tracking, so there was no opportunity to analyse the potential impacts of eye movements on their behaviour. We also did not instruct the participants to prioritise the speed of their response, so we do not know whether there was a relationship between the imbalance and the speed with which the task could be performed. These aspects may be of scientific interest in future investigations.

## Data Availability

All data produced in the present study are available upon reasonable request to the authors.

## 5 Additional information

### 5.1 Acknowledgements

The authors thank MiYoung Kwon and Peter Bex for sharing the code of the Dichoptic Letters test. This pre-print manuscript was written in LATEX using TeXShop, and is formatted with a custom style available at: github.com/alexsbaldwin/biorxiv-inspired-latex-style.

### 5.2 Author contributions in CREDIT format

**Alex S Baldwin**: Conceptualization, Methodology, Software, Validation, Formal Analysis, Data Curation, Writing - Original Draft, Writing - Review & Editing, Visualization, Supervision, Project Administration, Funding Acquisition; **Marie-Céline Lorenzini**: Investigation, Writing - Review & Editing; **Annabel Wing-Yan Fan**: Investigation, Software, Validation, Writing - Review & Editing; **Robert F Hess**: Conceptualization, Resources, Writing - Original Draft, Writing Review & Editing, Supervision, Project Administration, Funding Acquisition; **Alexandre Reynaud**: Conceptualization, Investigation, Writing - Original Draft, Writing - Review & Editing, Supervision, Project Administration, Funding Acquisition.

## 6 Appendix

### 6.1 Table of Suppression Measurement Methods

The findings from a review of previously-published methods for measuring binocular imbalance (such as from interocular suppression) are presented in Table A1.

### 6.2 Algorithm Details

The goal of the measurement is to estimate the underlying interocular contrast ratio that defines the imbalance in the perceived contrast between the eyes. This is found as the most likely value of the interocular ratio parameter (*r*) based on the history of ranking responses given to the different stimulus conditions

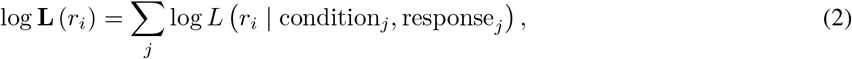

where log **L** (*r*_*i*_) is the overall log-likelihood of the *i*^th^ parameter value, calculated as the sum of the log-likelihoods based on the responses given to the conditions presented over each *j* of the trials. These likelihood values are obtained from pre-generated look-up tables. After converting from log-likelihood back to likelihood, we normalise to give **L**^*′*^

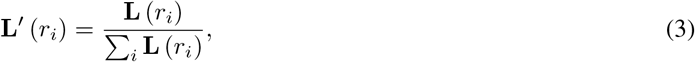

where the sum of **L**^*′*^ over each *i* of the possible ratios is 1. This **L**^*′*^ (*r*_*i*_) is our running estimate of the likelihood of each of our discretely-sampled parameter values *r*_*i*_. The algorithm attempts to estimate the most likely of these parameter values. This is done by minimising the expected posterior entropy. The entropy given all previous trials is defined as

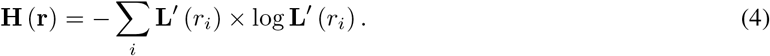

To obtain the expected posterior entropy, we must consider adding one more trial and account for the various possible responses that the participant is more or less probable to give based on the current best estimate of their interocular ratio. To find the expected entropy resulting from the *j* + 1 trial, we determine the normalised likelihood function that would be the outcome for each possible ranking response in that trial. We use Bayes’ Theorem, simply

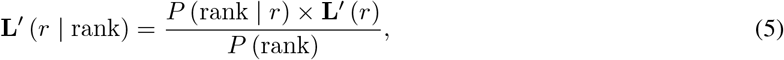

where **L**^*′*^ (*r*) is the prior likelihood of the ratio value, and *P* (rank) is the marginal probability of the ranking outcome. This probability, *P* (rank), of giving a certain (*k*^th^) ranking response has to take into account the relative likelihoods of the different interocular ratio parameter options

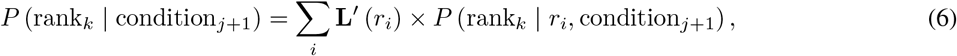

where the probability of a certain ranking being given to a particular condition under a particular interocular ratio parameter, *P* (rank_*k*_ | *r*_*i*_, condition_*j*+1_), can be obtained directly from the Monte-Carlo generated look-up tables.

**Table A1:** Results of a review of existing methods for measuring interocular suppression.

| Relevant study | Identifier | Parameter measured | Potential disadvantage | Special contribution |
| --- | --- | --- | --- | --- |
| Wolfe (1986) | Binocular rivalry | Duration | Artificial viewing condition | Dynamic nature |
| Hess (1991) | Contrast adaptation | Contrast | Decays over time | Putative site in V1 |
| Harrad and Hess (1992) | Dichoptic increment detection | Contrast |  | Contrast-level dependence |
| Tsuchiya et al. (2006) | Continuous flash suppression | Contrast |  | Extended suppression of stimulus from awareness |
| Huang et al. (2009) | Phase combination | Contrast / Phase | Limited to lower spatial frequencies | Measures balance in fusion state |
| Black et al. (2011) | Global motion | Contrast / Motion | Time-consuming | Putative site in extra-striate cortex |
| Li et al. (2011) | Bagolini striated glass | Contrast | Uncontrolled room lighting |  |
| Maehara et al. (2011) | Contrast combination | Contrast |  | Spatial frequency dependence |
| Huang et al. (2012) | Acuity measure | Contrast |  | Spatio-temporal properties |
| Babu et al. (2013) | Regional contrast matching | Contrast | Local | Measures regional visual field dependence |
| Zhou et al. (2013) | Global orientation | Contrast / Orientation |  | Putative site in extra-striate cortex |
| Kwon et al. (2015) | Dichoptic letter test | Contrast | Measurement under artificial dichoptic mismatch. | Rivalry-based (winner take all). Acuity dependence. |
| Spiegel et al. (2016) | Blurred edge combination | Contrast / Position | Time-consuming. | Highlights the probabilistic nature of suppression. |
| Bossi et al. (2017) | Dichoptic increment/decrement | Contrast | Increment/decrement nonlinearity presented dichoptically | Performed while watching movies |
| Zhou et al. (2018) | Noise masking | Contrast | Off-frequency effects | Spatial frequency dependence |
| Wang et al. (2019) | Orientation combination | Contrast / Orientation | Cross-orientation effects | Spatial frequency dependence of fusional consequences |
| Webber et al. (2020) | Quantitative Worth 4 Dot | Contrast | Colour-contrast effects | Relevance to existing clinical tool |

Substituting the appropriate values into Equation 5, we have

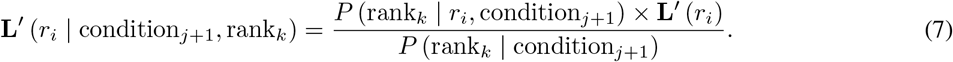

The conditional entropy of the likelihood function from that single trial outcome (a specific ranking response given to the presented condition) is then calculated

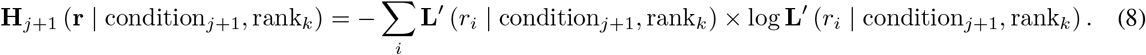

The expected entropy resulting from the *j* + 1 trial given a certain condition is the weighted average of the entropies calculated from the likelihood functions for each *k* of the different possible ranking responses. They are weighted by the probability of each ranking response being given

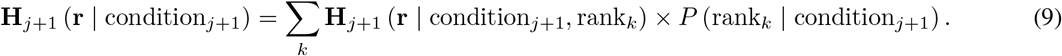

**Figure A1:**
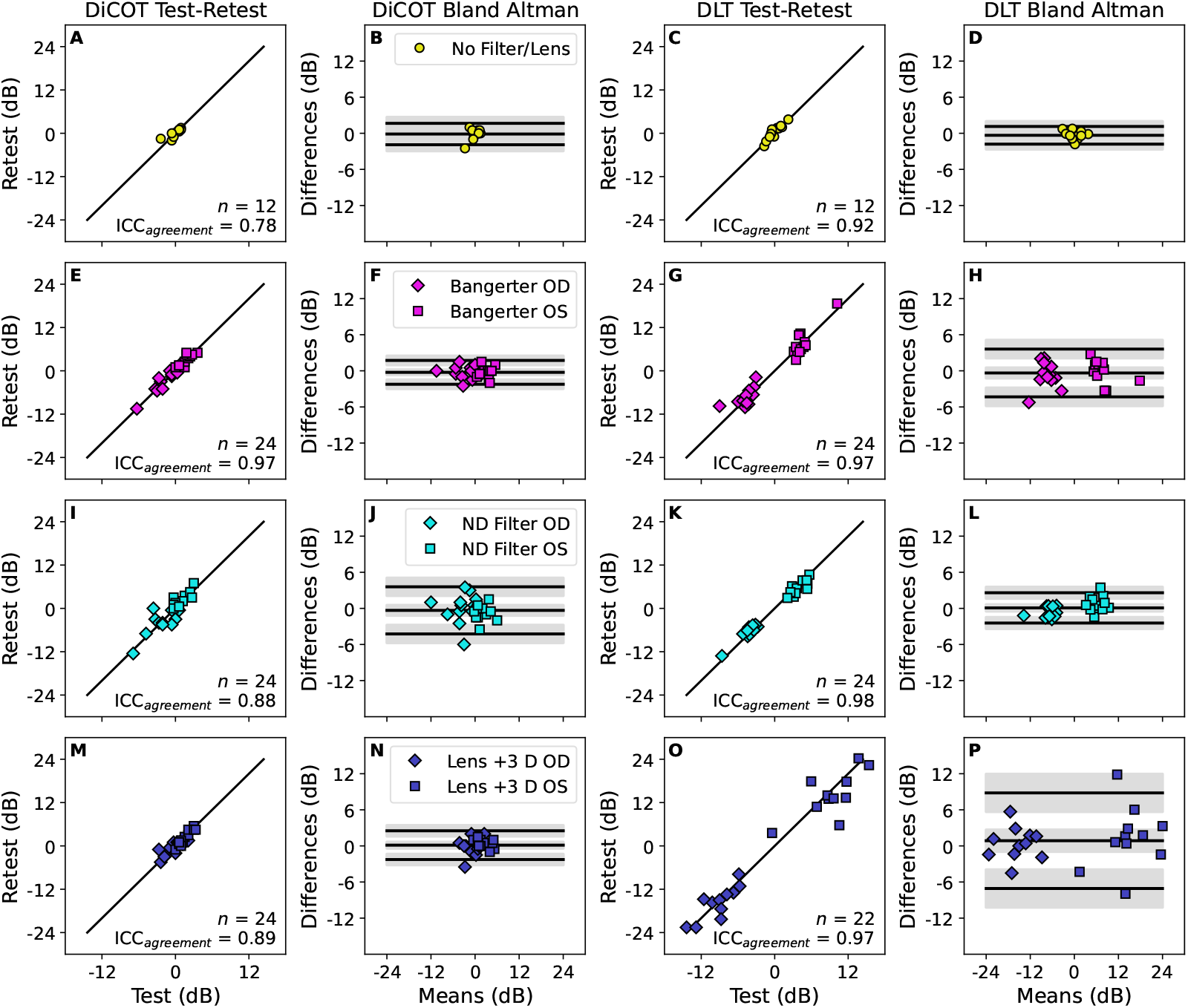
Test-retest reliability. The data of Figure 4 are replotted here, separated into the different filter or lens conditions (rows). The first two columns show the DiCOT measurements. The remaining two columns show data from the DLT. In each pair, the left column shows correlations between test and retest measurements. The solid diagonal line indicates equality. Each point indicates a data point from one participant in one lens/filter condition. The right column shows Bland-Altman plots giving the test-retest difference as a function of the mean of the two measures. The three black lines show the bias and the upper and lower limits of agreement. The shaded region around each line gives the 95% confidence interval.

The conditions for the next trial (condition_*j*+1_) are therefore chosen to minimise the value of Equation 9. As we consider only a relatively small set of possible conditions, we are able to calculate this value for all of the candidates and select the one that produces the smallest value.

### 6.3 Supplementary Analysis

In addition to the combined analysis given in Figure 4, we also analysed the test-retest data for each individual filter or lens condition. This is plotted in Figure A1, with the statistics for these individual conditions reported in Table 1.

When comparing the two measurement tools (DLT vs DiCOT), we also analysed the data from the individual conditions separately. This is presented in Figure A2. The regression slopes from these plots are summarised in Table 2, with the complete analysis reported in Table A1. A significant linear relationship is found between the DLT and DiCOT results for all conditions except “No Filter/Lens”, due to the relatively small variation in binocular imbalance in our population. As in Figure 5, because the slope of the relationship in the scatter plots is not unity, we find a strong relationship between the DLT result value and the DLT-DiCOT error in the Bland-Altman analysis (where the slope of that relationship is equal to one minus the slope from the scatter plot).

**Figure A2:**
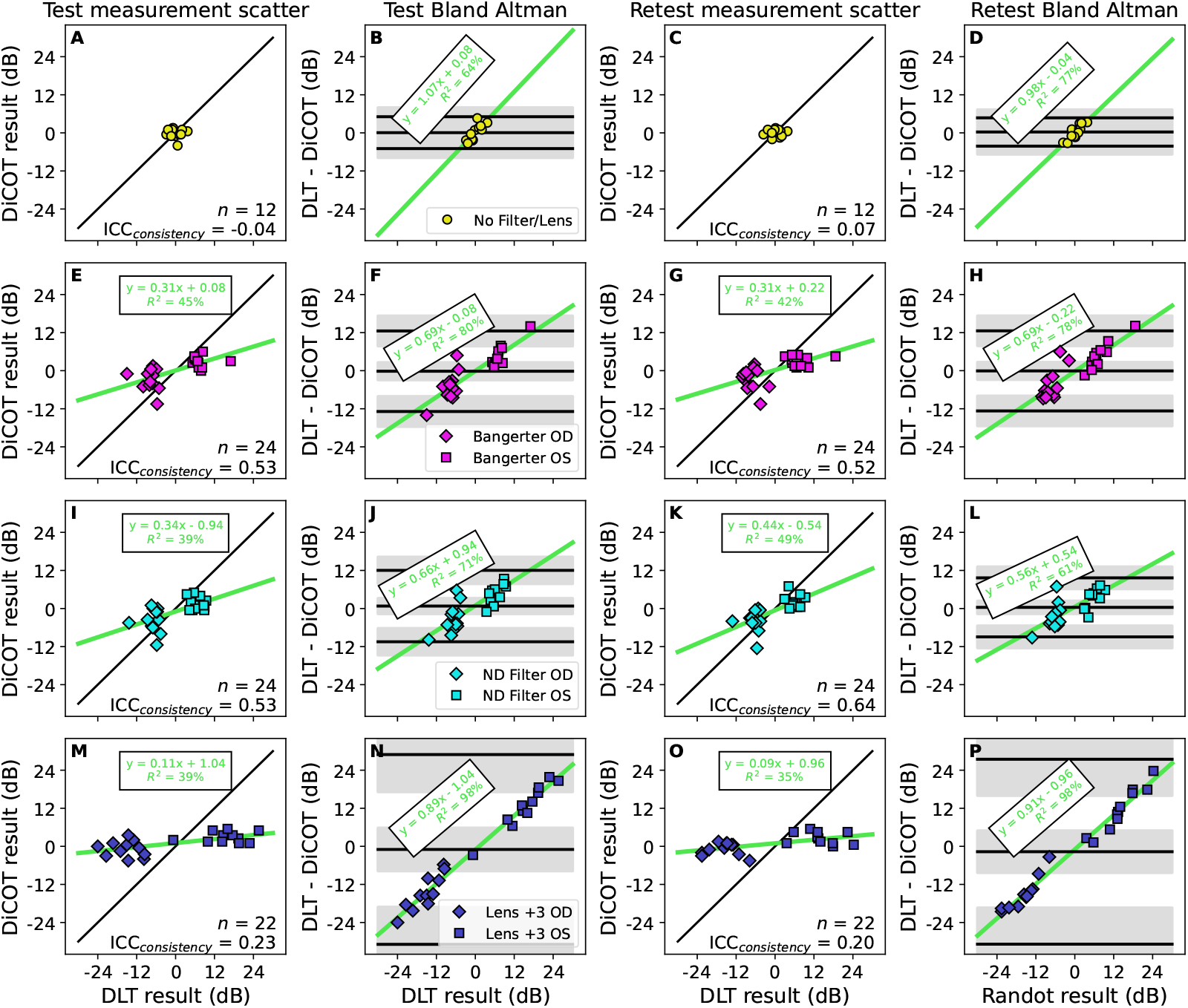
Comparison between DLT and DiCOT. The data of Figure 5 are replotted here, separated into the different filter or lens conditions (rows). The first two columns show comparisons of the test (first) measurements. The remaining two columns show comparisons of the retest (second) measurements. In each pair, the left column shows scatterplots comparing the measurement made with one test against that made with the other. The diagonal green line is the best-fitting linear relationship (parameters given in green text). The black line shows equality. The right column shows Bland-Altman plots derived from the scatter plots. The three black lines show the bias and the upper and lower limits of agreement. The shaded region around each line gives the 95% confidence interval. The diagonal green line is the best-fitting linear relationship (parameters given in green text).

In Figure 6, we transformed the DiCOT results according to the overall linear relationship found with the DLT data. Here, we report an additional analysis where this transformation was performed separately for each filter or lens condition (n.b. the “No Filter/Lens” condition is excluded as there was no significant relationship to base the transformation on). This analysis is shown in Figure A3, with statistics reported in Table A3. The transformation accounts for both the slope and bias of the relationships reported in the scatter plots in Figure A2. Therefore, after the transformation the scatter plot relationship is, by definition, *y* = *x*. There is no bias on the Bland-Altman plot and the slope of the regression in that plot is zero. The ICC_consistency_ score is improved by this adjustment, though there remains a wide limits of agreement range on the Bland-Altman plot.

**Table A2:**
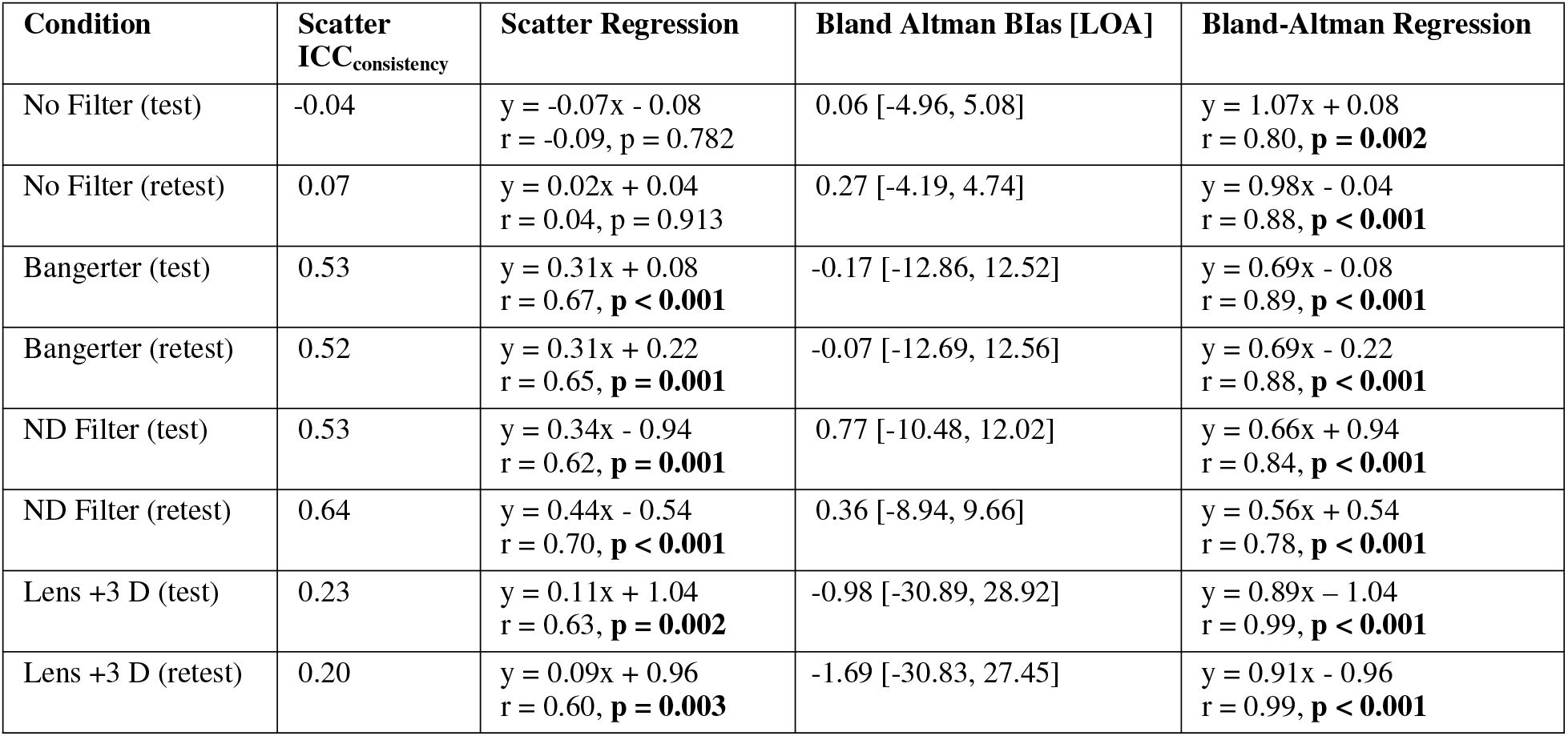
Comparison between DLT and DiCOT. Statistics from Figure A2, giving the consistency form of the Intraclass Correlation Coefficient, as well as the linear regression analysis, the bias and limits of agreement from the Bland-Altman plots, and the linear regression analysis from the Bland-Altman plots.

**Figure A3:**
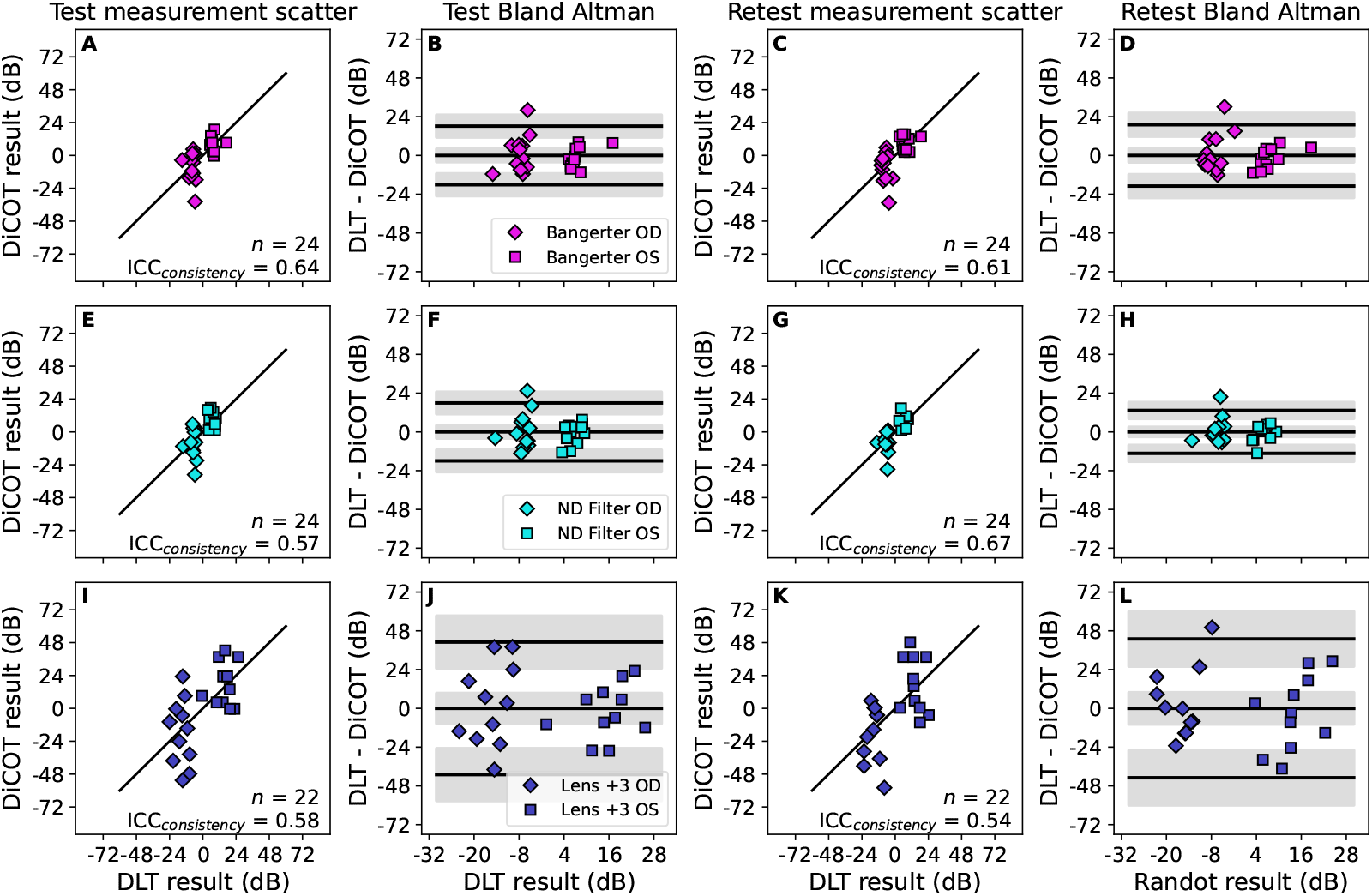
Comparison between DLT and adjusted DiCOT measurements. Similar to Figure A2, but with DiCOT values adjusted according to the linear relationships reported in Figure A2.

**Table A3:**
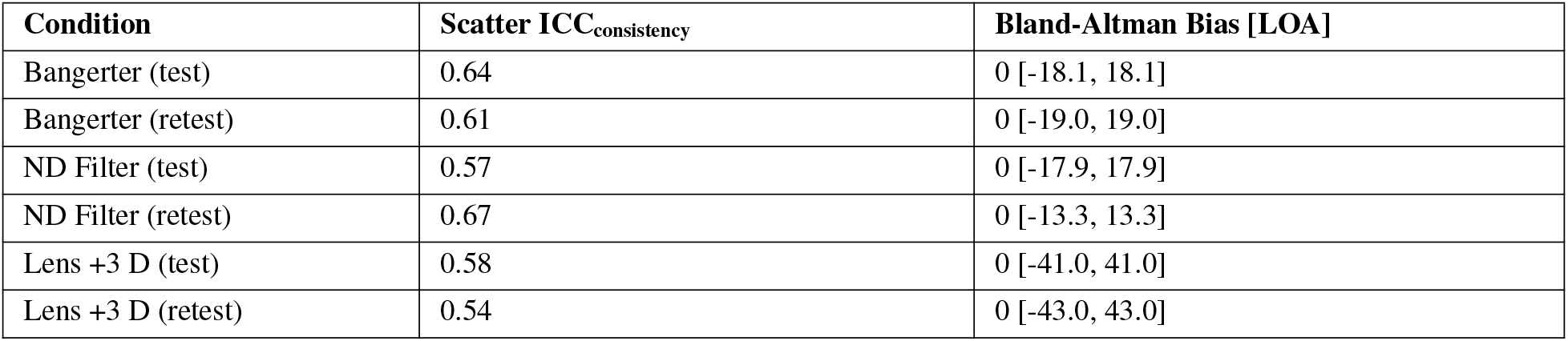
Comparison between DLT and adjusted DiCOT measurements. Statistics from Figure A3, giving the consistency form of the Intraclass Correlation Coefficient, as well as the bias and the limits of agreement from the Bland-Altman plots.

